# Adjuvant Trastuzumab Deruxtecan Versus Trastuzumab Emtansine by Residual Cancer Burden in HER2-Positive Early Breast Cancer: A Decision Analysis

**DOI:** 10.64898/2026.10.19.26365942

**Authors:** Jane Fan, Axel Stenmark Tullberg, Allison W. Kurian, Jennifer L. Caswell-Jin, James C. Dickerson, Fernando Alarid-Escudero

**Affiliations:** Department of Epidemiology and Population Health, Stanford University School of Medicine, Stanford, CA; Department of Medicine (Oncology), Stanford University School of Medicine, Stanford, CA; Department of Health Policy, Stanford University School of Medicine, Stanford, CA; Center for Health Policy, Freeman Spogli Institute for International Studies, Stanford University, Stanford, CA

**Keywords:** breast cancer, adjuvant, trastuzumab deruxtecan, trastuzumab emtansine, survival

## Abstract

**Background:** For patients with HER2-positive early breast cancer and residual disease after neoadjuvant therapy, escalating adjuvant treatment improves outcomes. In high-risk patients, DESTINY-Breast05 found that trastuzumab deruxtecan (T-DXd) reduced the 3-year risk of recurrence compared with trastuzumab emtansine (T-DM1). T-DXd also carries a risk of fatal interstitial lung disease (ILD). The US FDA approval for adjuvant T-DXd includes all patients with residual disease, including lower-risk patients not represented in the trial. We used decision modeling to project 10-year survival across recurrence-risk groups to aid in clinical decision-making.

**Methods:** We modeled outcomes for 50-year-old patients receiving T-DXd or T-DM1 using DESTINY-Breast05 data and published literature. The primary outcome was projected 10-year overall survival (OS); secondary outcomes were breast cancer and ILD deaths per 1,000 patients. Results were stratified by residual cancer burden (RCB) class and estrogen receptor (ER) status.

**Results:** Across 1,000 simulations, mean projected OS differences favored T-DXd in every subgroup, with 10-year gains ranging from 0.3 (95% uncertainty interval [UI], -2.1 to 1.6; RCB-I ER-positive) to 9.7 percentage points (95% UI, 5.7 to 14.1; RCB-III ER-positive). T-DXd yielded higher OS in 71% of RCB-I ER-positive simulations and 94% of RCB-I ER-negative simulations. Per 1,000 patients, T-DXd averted 16 to 113 breast cancer deaths and caused 13 ILD deaths compared with T-DM1, assuming a conservative fatal ILD risk.

**Conclusion:** These findings favor T-DXd for patients with RCB-II or RCB-III disease. For RCB-I patients, particularly those with ER-positive tumors, the projected survival gain is smaller and sensitive to fatal ILD assumptions.

## Introduction

The standard of care for stage II to III HER2-positive breast cancer is preoperative chemotherapy with HER2-targeted therapy, with the goal of achieving a pathologic complete response (pCR) at surgery. However, approximately 40% to 60% of patients have residual invasive disease, which is associated with a higher risk of recurrence and death.^1^ For these patients, the KATHERINE trial demonstrated that escalating adjuvant treatment from trastuzumab to the antibody-drug conjugate trastuzumab emtansine (T-DM1) improves overall survival (OS). Following its approval by the US Food and Drug Administration (FDA) in 2019, T-DM1 became the standard adjuvant treatment for patients with residual disease.

In 2025, the DESTINY-Breast05 trial showed that adjuvant trastuzumab deruxtecan (T-DXd) reduced the 3-year risk of invasive recurrence or death compared with T-DM1.^2^ Eligibility was restricted to patients with residual node-positive disease or disease that was inoperable at presentation, who represent approximately half of patients with residual disease.^3^ In May 2026, the FDA approved adjuvant T-DXd for patients “with residual invasive disease”,^4^ an indication substantially broader than the high-risk trial population.

In the curative-intent setting, the risk of fatal interstitial lung disease (ILD) with T-DXd must be justified by a meaningful reduction in breast cancer deaths.^5,6^ For lower-risk patients covered by the FDA label but not represented in the trial, the absolute reduction in recurrence is expected to be smaller while the risk of fatal ILD remains, creating uncertainty about the net survival benefit in these patients. Recent commentaries have proposed tailoring escalation by residual disease,^5,7^ but there are no quantitative estimates of benefit and harm for lower-risk patients. We are unaware of any planned randomized trial of T-DXd in this population, and observational survival data will take years to mature, yet oncologists are facing this treatment decision in clinical practice now. Decision analysis is well-suited to address this problem: rather than predicting outcomes for an individual patient, it allows the randomized comparison from DESTINY-Breast05 to be combined with external evidence on baseline recurrence risk to compare the expected benefits and harms of each treatment under explicit assumptions that can be tested. We used this approach to estimate whether the expected reduction in breast cancer deaths with T-DXd outweighs its risk of fatal ILD across levels of residual disease, and how high the ILD risk would need to be to favor T-DM1 instead.

To make our results clinically interpretable, we quantified the risk of recurrence based on residual disease and estrogen receptor (ER) status. We used the residual cancer burden (RCB) index, which measures the extent of disease remaining in the breast and axillary nodes and is categorized into four classes, from RCB-0 (pCR) to RCB-III (extensive residual disease).^8,9^ The RCB class is a common reporting measure on pathology reports and is prognostic in HER2-positive breast cancer, with the risk of recurrence increasing across classes.^9,10^ ER status also influences outcomes, with ER-positive HER2-positive cancers associated with lower rates of pCR and prolonged patterns of recurrence relative to ER-negative cancers.^11-14^ We present our results stratified across RCB class and ER status to examine how the projected survival difference between T-DXd and T-DM1 varies across these groups.

## Methods

### Model Assumptions

We developed a state-transition decision-analytic model to compare clinical outcomes with adjuvant T-DXd versus T-DM1 in a hypothetical cohort of 50-year-old patients with HER2-positive early breast cancer and residual invasive disease after neoadjuvant therapy over a 10-year simulation horizon (**Figure 1**). Age 50 was chosen to be consistent with the median age in DESTINY-Breast05^2^ and other decision-modeling studies in HER2-positive early breast cancer.^15-17^

**Figure 1.**
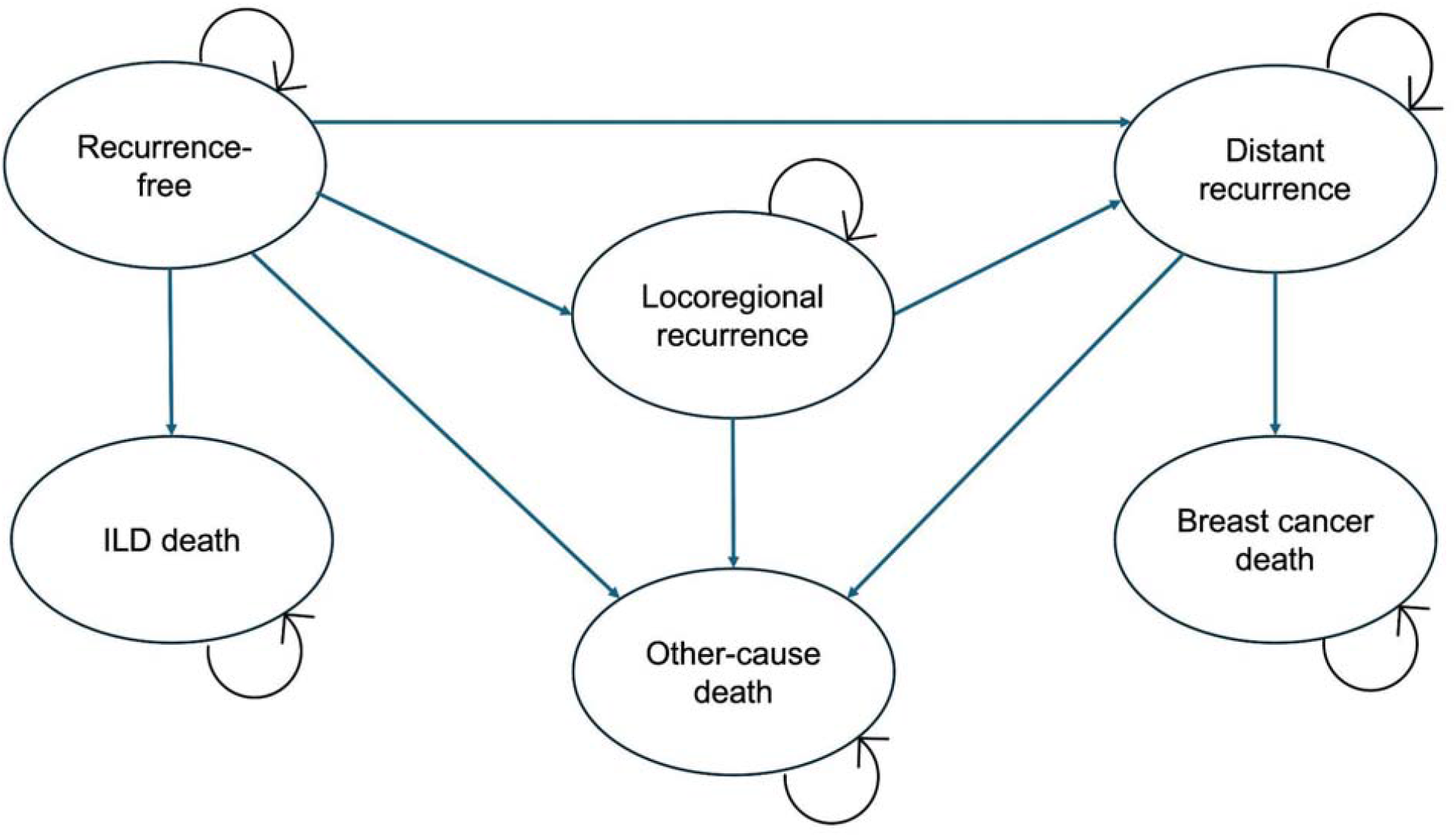
Cohort State-Transition Diagram of the Clinical Decision-Analytic Model^a^. Abbreviations: ILD, interstitial lung disease. ^a^ Shows the states and transitions used in the model. For visual clarity, the recurrence-free state aggregates the treatment-specific on-treatment, off-treatment, and month-indexed post-ILD discontinuation states. Treatment discontinuation and switching transitions are not shown. Death from ILD can occur only during adjuvant treatment. All non-death states are subject to other-cause mortality. All death states are absorbing.

We made several key assumptions in our model. All patients entered the model recurrence-free and were assigned to adjuvant T-DXd or T-DM1 for a 10-month treatment period, consistent with standard dosing every 3 weeks for a total of 14 treatment cycles.^2^ The model used monthly cycles, during which patients could remain recurrence-free, develop locoregional or distant recurrence, or die. The risk of recurrence depended on their RCB class and ER status. Patients who experienced locoregional recurrence could progress to distant recurrence at any time after their initial recurrence. Those with distant recurrence faced an additional risk of death from breast cancer. We assumed that all ILD events occurred while patients were on their adjuvant therapy. Based on clinical practice and the DESTINY-Breast05 protocol for ILD management, we modeled grades 2 to 4 ILD as resulting in permanent treatment discontinuation. Patients who discontinued T-DXd received an *a priori*-specified fraction of the full treatment effect based on the month of discontinuation, with diminishing incremental gains in treatment benefit over time and the full benefit attained for patients who received T-DXd for the full 10 months. We assumed that T-DM1 was started in the cycle following T-DXd discontinuation. Given that no fatal ILD cases were reported for T-DM1 in DESTINY-Breast05, we did not model fatal ILD for T-DM1. We also did not account for contralateral invasive breast cancer due to its low frequency in DESTINY-Breast05.

### Model Inputs and Calibration

Model inputs were informed by DESTINY-Breast05 and other relevant data sources (**Table 1**). DESTINY-Breast05, the only randomized comparison of T-DXd and T-DM1 in HER2-positive early breast cancer, informed the relative treatment effect, ILD risk, and calibration targets. The Yau et al pooled analysis informed baseline recurrence risk by RCB class and ER status,^10^ which we adjusted for adjuvant T-DM1 using the final analysis of the KATHERINE trial.^18^ US life tables informed other-cause mortality.^19^ Disease progression parameters were obtained by calibrating the model to the 3-year overall survival (OS), invasive disease-free survival (iDFS), and distant recurrence-free interval (DRFI) endpoints reported in DESTINY-Breast05, as well as to the approximate proportion of recurrences that were locoregional across both arms (∼10%), based on Table S2 from DESTINY-Breast05.^2^ We note that DESTINY-Breast05 reported fatal ILD in 0.25% of patients assigned to T-DXd,^2^ which was used in model calibration. However, because trial eligibility and protocol-mandated monitoring may underestimate real-world toxicity,^20^ our primary analysis following calibration incorporated a 1.0% probability of fatal ILD among patients assigned to T-DXd, drawing on pooled T-DXd safety analyses^21,22^ and clinical expertise. This conservative fatal ILD assumption favors T-DM1 and strengthens the robustness of findings favoring T-DXd; however, it substantially influences the magnitude of the projected benefit in RCB-I disease. As such, we also report results informed by the trial-observed fatal ILD risk. We accounted for potential non-identifiability in calibration and uncertainty in the calibrated parameters by adopting a Bayesian approach using the Incremental Mixture Importance Sampling algorithm.^23^ Given the lack of long-term follow-up data regarding the persistence of the T-DXd treatment effect in this population, we modeled the T-DXd hazard ratio for distant recurrence as waning linearly after year 3 (the time point at which efficacy outcomes were reported in DESTINY-Breast05) to no effect (hazard ratio = 1) until year 10 as a plausible but conservative estimate of T-DXd benefit durability. Additional details on model inputs and calibration are available in the Supplementary Materials (**Supplementary Methods; Figures S1-S3; Table S1**).

**Table 1.**
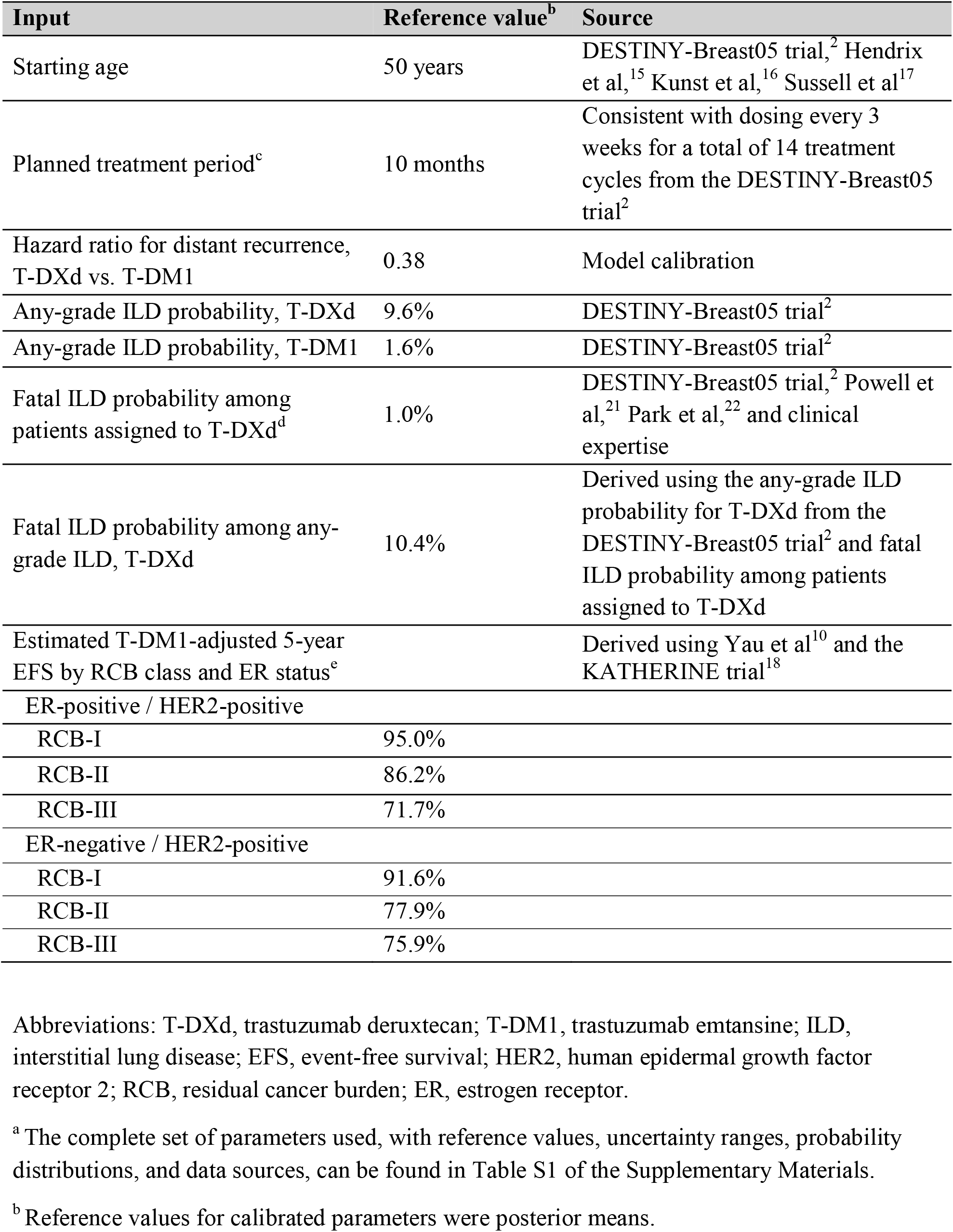
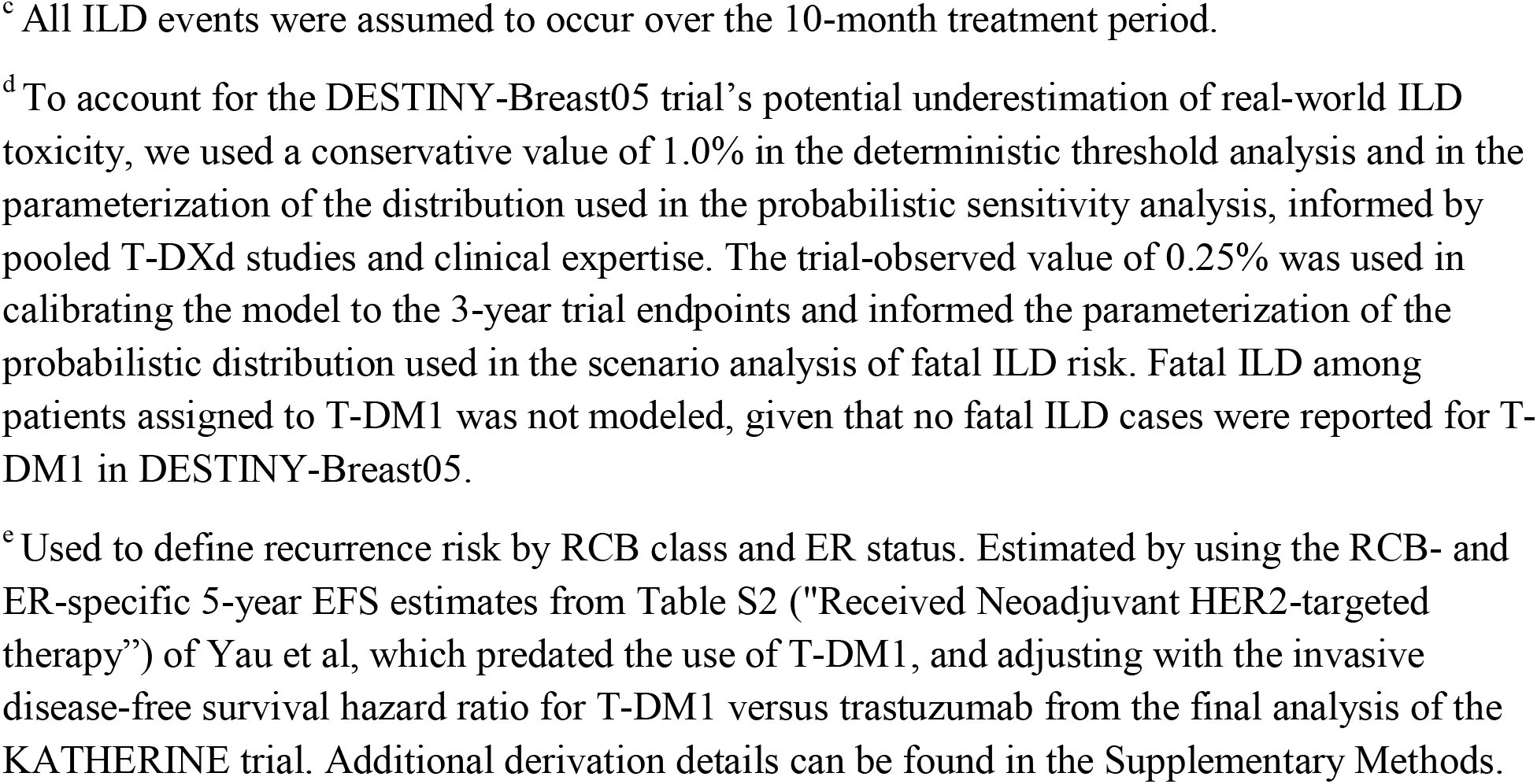
Select Parameters for the Clinical Decision-Analytic Model^a^.

| Input | Reference value<sup>b</sup> | Source |
| --- | --- | --- |
| Starting age | 50 years | DESTINY-Breast05 trial,<sup>2</sup> Hendrix et al,<sup>15</sup> Kunst et al,<sup>16</sup> Sussell et al<sup>17</sup> |
| Planned treatment period<sup>c</sup> | 10 months | Consistent with dosing every 3 weeks for a total of 14 treatment cycles from the DESTINY-Breast05 trial<sup>2</sup> |
| Hazard ratio for distant recurrence, T-DXd vs. T-DM1 | 0.38 | Model calibration |
| Any-grade ILD probability, T-DXd | 9.6% | DESTINY-Breast05 trial<sup>2</sup> |
| Any-grade ILD probability, T-DM1 | 1.6% | DESTINY-Breast05 trial<sup>2</sup> |
| Fatal ILD probability among patients assigned to T-DXd<sup>d</sup> | 1.0% | DESTINY-Breast05 trial,<sup>2</sup> Powell et al,<sup>21</sup> Park et al,<sup>22</sup> and clinical expertise |
| Fatal ILD probability among any-grade ILD, T-DXd | 10.4% | Derived using the any-grade ILD probability for T-DXd from the DESTINY-Breast05 trial<sup>2</sup> and fatal ILD probability among patients assigned to T-DXd |
| Estimated T-DM1-adjusted 5-year EFS by RCB class and ER status<sup>e</sup> |  | Derived using Yau et al<sup>10</sup> and the KATHERINE trial<sup>18</sup> |
| ER-positive / HER2-positive |  |  |
| RCB-I | 95.0% |  |
| RCB-II | 86.2% |  |
| RCB-III | 71.7% |  |
| ER-negative / HER2-positive |  |  |
| RCB-I | 91.6% |  |
| RCB-II | 77.9% |  |
| RCB-III | 75.9% |  |

Abbreviations: T-DXd, trastuzumab deruxtecan; T-DM1, trastuzumab emtansine; ILD, interstitial lung disease; EFS, event-free survival; HER2, human epidermal growth factor receptor 2; RCB, residual cancer burden; ER, estrogen receptor.
<sup>a</sup> The complete set of parameters used, with reference values, uncertainty ranges, probability distributions, and data sources, can be found in Table S1 of the Supplementary Materials.
<sup>b</sup> Reference values for calibrated parameters were posterior means.
<sup>c</sup> All ILD events were assumed to occur over the 10-month treatment period.
<sup>d</sup> To account for the DESTINY-Breast05 trial's potential underestimation of real-world ILD toxicity, we used a conservative value of 1.0% in the deterministic threshold analysis and in the parameterization of the distribution used in the probabilistic sensitivity analysis, informed by pooled T-DXd studies and clinical expertise. The trial-observed value of 0.25% was used in calibrating the model to the 3-year trial endpoints and informed the parameterization of the probabilistic distribution used in the scenario analysis of fatal ILD risk. Fatal ILD among patients assigned to T-DM1 was not modeled, given that no fatal ILD cases were reported for T-DM1 in DESTINY-Breast05.
<sup>e</sup> Used to define recurrence risk by RCB class and ER status. Estimated by using the RCB- and ER-specific 5-year EFS estimates from Table S2 ("Received Neoadjuvant HER2-targeted therapy") of Yau et al, which predated the use of T-DM1, and adjusting with the invasive disease-free survival hazard ratio for T-DM1 versus trastuzumab from the final analysis of the KATHERINE trial. Additional derivation details can be found in the Supplementary Methods.

### Outcomes

The primary outcome was the difference in projected 10-year OS between T-DXd and T-DM1, in percentage points. Secondary outcomes included breast cancer deaths averted and ILD deaths caused per 1,000 patients, with T-DXd versus T-DM1, over the 10-year simulation horizon.

### Statistical Analysis

We conducted a probabilistic analysis, propagating parameter uncertainty across 1,000 draws and reporting means with 95% uncertainty intervals (UIs). In the primary analysis, we assumed linear waning of the benefit of T-DXd relative to T-DM1 after year 3 until year 10 and a conservative fatal ILD risk. With these assumptions, we evaluated the primary outcome of 10-year OS under a base case and three edge cases: (i) an attenuated T-DXd hazard ratio for distant recurrence of 0.80; (ii) doubled probability of any-grade ILD with T-DXd, with a proportional increase in the probability of fatal ILD among patients assigned to T-DXd; and (iii) both combined. For each edge case, the specified transformation was applied to the relevant sampled parameter values, while all other sampled inputs were left unchanged. We evaluated secondary outcomes probabilistically under the base case only. We also conducted a one-way deterministic threshold analysis under base-case assumptions to estimate how much T-DXd-associated ILD is needed to offset the projected survival improvement from reduced breast cancer deaths.

As our assumption about the persistence of the T-DXd treatment effect beyond the 3-year timepoint at which efficacy outcomes were reported in DESTINY-Breast05 may be critical to model outputs, we performed a structural sensitivity analysis. We repeated the base-case probabilistic analysis of the primary outcome under two alternative treatment-effect persistence assumptions: (i) cessation of T-DXd benefit immediately after year 3, with the hazard ratio reverting to 1; and (ii) persistent (constant) benefit through year 10. In each structural scenario, all other parameters remained unchanged.

Lastly, because the conservative T-DXd-associated fatal ILD assumption favors T-DM1 and may materially affect the magnitude, and potentially the direction, of projected OS differences in RCB-I disease in addition to the magnitude of differences in ILD deaths, we conducted a scenario analysis in which we repeated the base-case probabilistic analysis for the primary and secondary outcomes with the distribution of fatal ILD among patients assigned to T-DXd informed by ILD event counts observed in DESTINY-Breast05, leaving all other sampled inputs unchanged. We additionally evaluated the primary outcome under the alternative treatment-effect persistence assumptions. All analyses were performed using R version 4.3.2 and the code is publicly available.

## Results

### Probabilistic Base Case Analysis

Across 1,000 base-case simulations under the primary model assumptions of treatment-effect waning and a conservative fatal ILD risk, mean projected OS differences favored T-DXd across all RCB and ER subgroups, although benefit was smallest and least certain for RCB-I ER-positive disease (**Table 2**). In ER-positive disease, the projected OS gain with T-DXd was 0.3 (95% UI: -2.1 to 1.6), 4.3 (95% UI: 1.3 to 6.8), and 9.7 (95% UI: 5.7 to 14.1) percentage points for RCB-I, RCB-II, and RCB-III, respectively; in ER-negative disease, the projected OS gain was 1.9 (95% UI: -0.6 to 3.7), 7.6 (95% UI: 4.2 to 11.2), and 8.3 (95% UI: 4.7 to 12.2) percentage points, respectively. T-DXd had higher 10-year OS compared to T-DM1 in 71% and 94% of simulations in the RCB-I ER-positive and ER-negative subgroups, respectively (**Figure 2**). Across all RCB-II and RCB-III subgroups, T-DXd had higher 10-year OS relative to T-DM1 in 100% of simulations.

**Table 2.**
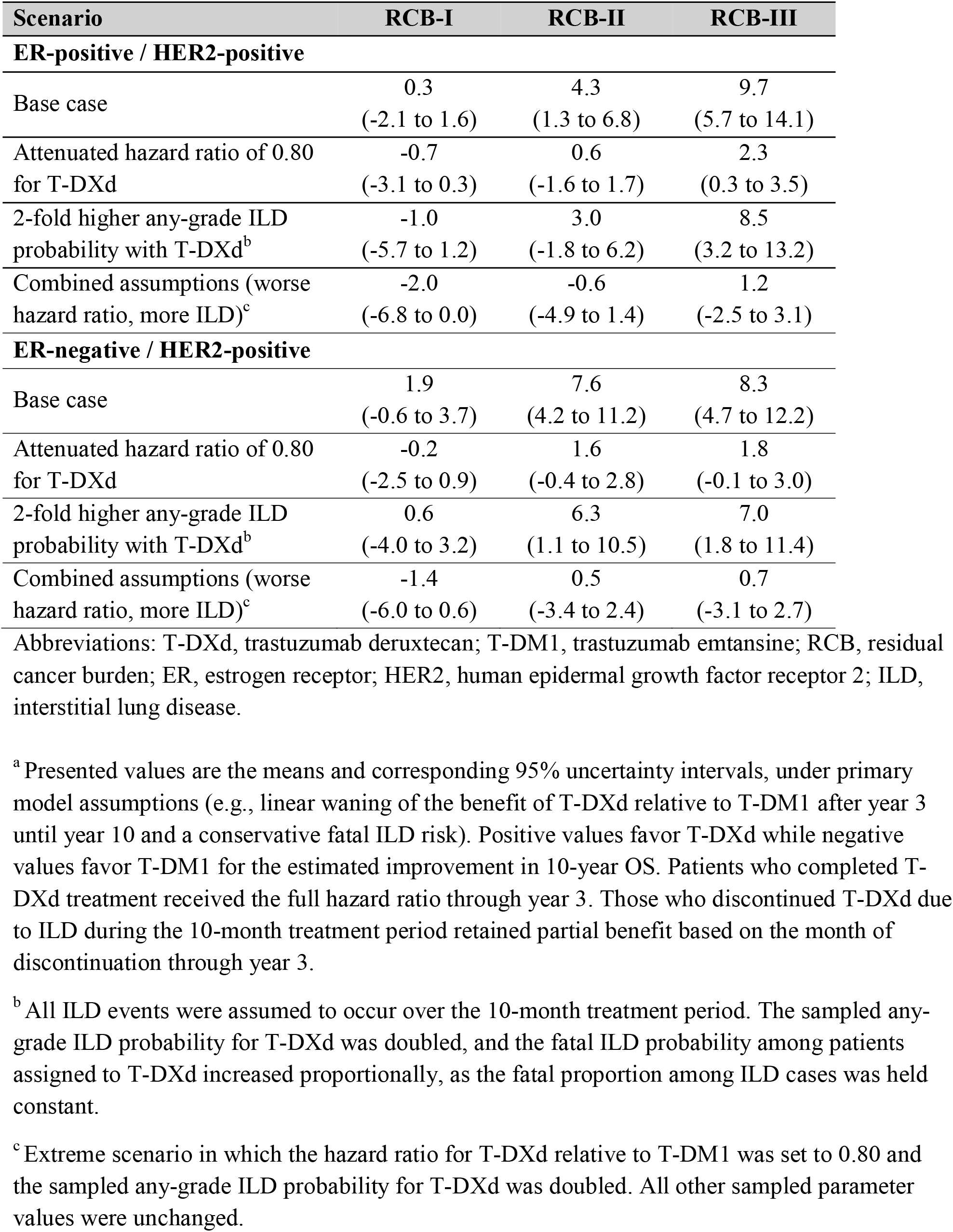
Mean Differences in 10-Year Overall Survival with T-DXd vs T-DM1 Across Base-Case and Edge-Case Scenarios^a^.

**Figure 2.**
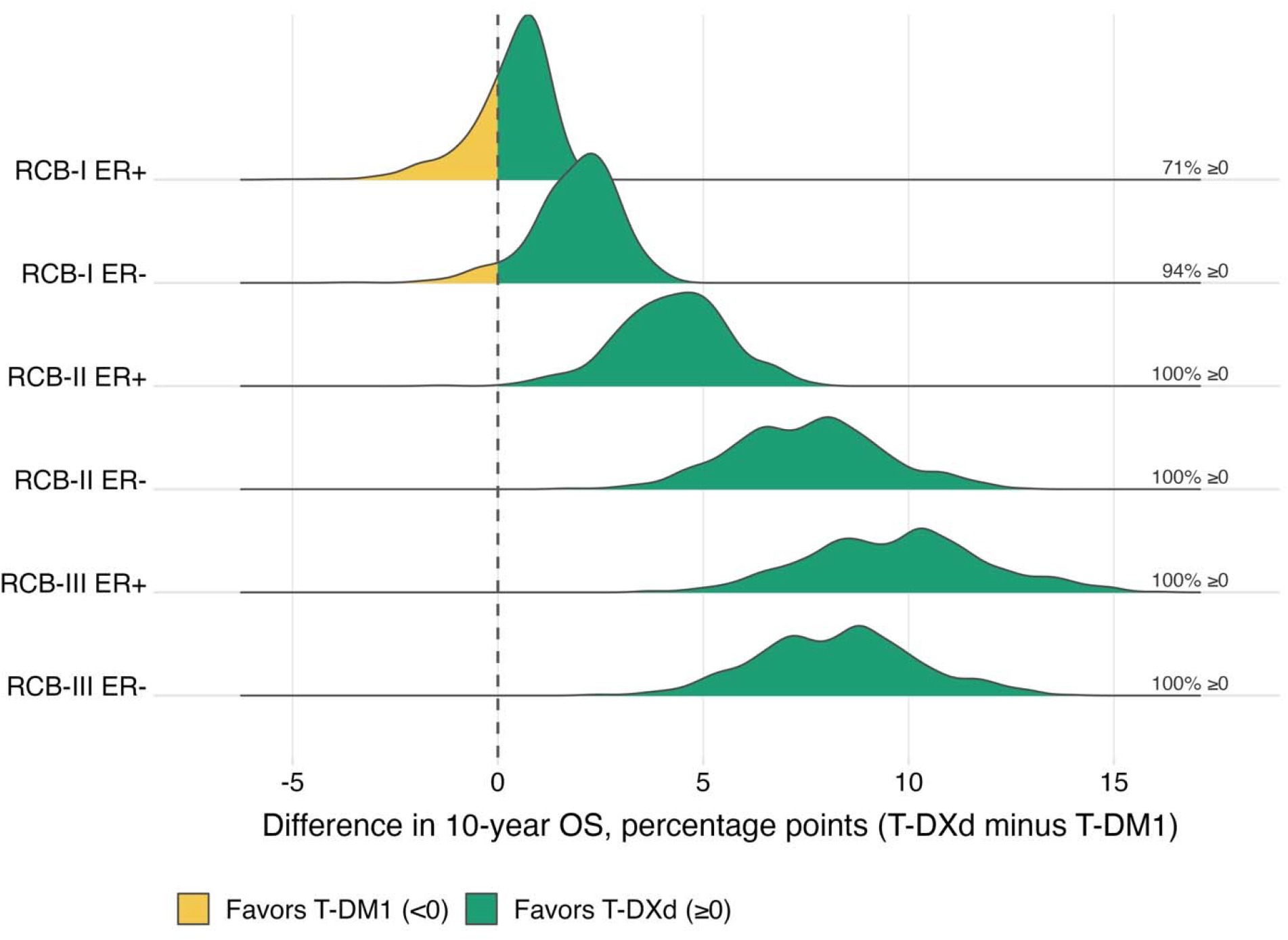
Probabilistic Distribution of the 10-Year Overall Survival Difference with T-DXd vs T-DM1^a^. Abbreviations: T-DXd, trastuzumab deruxtecan; T-DM1, trastuzumab emtansine; RCB, residual cancer burden; ER, estrogen receptor; OS, overall survival. ^a^ Shows the distribution of the absolute differences in 10-year OS with T-DXd vs T-DM1 across 1,000 simulations, reported in percentage points, under primary model assumptions (e.g., linear waning of the benefit of T-DXd relative to T-DM1 after year 3 until year 10 and a conservative fatal ILD risk). Positive values favor T-DXd while negative values favor T-DM1. The percentage of 1,000 simulations favoring T-DXd is shown to the right. Patients who completed T-DXd treatment received the full hazard ratio through year 3. Those who discontinued T-DXd due to ILD during the 10-month treatment period retained partial benefit based on the month of discontinuation through year 3.

Per 1,000 patients, T-DXd averted between 16.0 (RCB-I) and 112.7 (RCB-III) breast cancer deaths in ER-positive disease and between 32.3 (RCB-I) and 97.9 (RCB-III) breast cancer deaths in ER-negative disease (**Table 3**). It also caused 13.1 to 13.3 ILD deaths per 1,000 patients, a range consistent across subgroups, as ILD risk does not scale with recurrence risk.

**Table 3.** Mean Differences in 10-Year Secondary Outcomes with T-DXd vs T-DM1^a^.

| **Outcome** | **RCB-I** | **RCB-II** | **RCB-III** |
| --- | --- | --- | --- |
| **ER-positive / HER2-positive** |  |  |  |
| Breast cancer deaths averted per 1,000 patients | 16.0<br>(10.3 to 23.0) | 56.6<br>(36.6 to 80.1) | 112.7<br>(74.4 to 157.2) |
| ILD deaths caused per 1,000 patients | 13.3<br>(2.4 to 38.8) | 13.2<br>(2.4 to 38.6) | 13.1<br>(2.4 to 38.3) |
| **ER-negative / HER2-positive** |  |  |  |
| Breast cancer deaths averted per 1,000 patients | 32.3<br>(20.8 to 46.0) | 90.4<br>(59.3 to 126.6) | 97.9<br>(64.4 to 136.9) |
| ILD deaths caused per 1,000 patients | 13.3<br>(2.4 to 38.7) | 13.2<br>(2.4 to 38.5) | 13.2<br>(2.4 to 38.4) |

Abbreviations: T-DXd, trastuzumab deruxtecan; T-DM1, trastuzumab emtansine; RCB, residual cancer burden; ER, estrogen receptor; HER2, human epidermal growth factor receptor 2; ILD, interstitial lung disease.
<sup>a</sup> Presented values are the means and corresponding 95% uncertainty intervals across 1,000 simulations, under primary model assumptions (e.g., linear waning of the benefit of T-DXd relative to T-DM1 after year 3 until year 10 and a conservative fatal ILD risk). All differences were calculated as the outcome under T-DXd minus that under T-DM1, except for breast cancer deaths averted per 1,000 patients, which was calculated as breast cancer deaths under T-DM1 minus those under T-DXd.

### Probabilistic Edge Case Analysis

In the combined edge case, in which we assumed an attenuated hazard ratio and 2-fold higher any-grade ILD probability with T-DXd while other inputs remained the same as in the base case, the projected OS gain with T-DXd for RCB-I fell from 0.3 to -2.0 percentage points (ER-positive) and from 1.9 to -1.4 percentage points (ER-negative); the projected OS gain was -0.6 percentage points for RCB-II ER-positive disease (**Table 2**). The UIs crossed zero for RCB-II and RCB-III subgroups.

### Deterministic Threshold Analysis of ILD Risk

In the one-way deterministic threshold analysis under the primary model assumptions, T-DM1 became favored in RCB-I once the probability of any-grade ILD with T-DXd reached 16% (ER-positive) or 31% (ER-negative). For RCB-II and RCB-III, the thresholds for the T-DXd-associated ILD probability at which T-DM1 became favored were beyond clinically plausible values, ranging from 53% to 91%.

### Structural Sensitivity Analysis of Treatment Effect Persistence

Under the most conservative treatment-effect persistence assumption, in which T-DXd benefit ceased immediately after year 3, mean projected OS differences continued to favor T-DXd for the RCB-II and RCB-III subgroups but favored T-DM1 in RCB-I ER-positive disease (-0.4 percentage points; 95% UI, -2.7 to 0.7); T-DXd was favored in 40% and 77% of RCB-I ER-positive and ER-negative simulations, respectively (**Table S2; Figure S4**). Under the least conservative treatment-effect persistence assumption, in which the T-DXd benefit persisted through year 10, T-DXd was favored for RCB-I patients in 82% and 98% of simulations for ER-positive and ER-negative disease, respectively (**Figure S5**).

### Scenario Analysis of Fatal ILD Risk

Using a less conservative fatal ILD risk among patients assigned to T-DXd, informed by DESTINY-Breast05, the projected OS gain with T-DXd ranged from 1.2 (RCB-I ER-positive) to 10.5 (RCB-III ER-positive) percentage points under the primary waning treatment-effect assumption (**Table S3**). T-DXd had higher 10-year OS relative to T-DM1 in 100% of simulations across all subgroups under the primary and persistent effect assumptions; with effect cessation after year 3, T-DXd was favored in 95% of RCB-I ER-positive simulations and 100% of RCB-II and RCB-III simulations. Under the primary treatment-effect assumption, T-DXd averted between 15.9 (RCB-I) and 111.2 (RCB-III) breast cancer deaths in ER-positive disease and between 31.9 (RCB-I) and 96.6 (RCB-III) breast cancer deaths in ER-negative disease, per 1,000 patients, and caused 3.8 ILD deaths, with UIs spanning 0.7 to 9.4.

## Discussion

The volume of residual disease after neoadjuvant therapy is a strong patient-level predictor of recurrence risk in HER2-positive early breast cancer. DESTINY-Breast05 established T-DXd as the preferred adjuvant therapy for high-risk patients with residual node-positive disease (ypN1– ypN3) or residual disease following inoperable disease at presentation (cT4 or cN2–cN3),^2^ representing approximately half of patients with residual disease.^3^ The more challenging question is how to treat patients with lower-volume residual disease who fall within the FDA-labeled population but were not included in DESTINY-Breast05, given the risk of fatal ILD with T-DXd in the absence of prospective and real-world data. To our knowledge, this study is the first to quantify the long-term absolute survival benefit with T-DXd versus T-DM1 based on residual disease burden in HER2-positive early breast cancer, balancing treatment efficacy and toxicity.

Our decision model projected an improvement in 10-year OS with T-DXd relative to T-DM1, including among patients outside the DESTINY-Breast05 high-risk population. The clearest way to understand the observed differences across subgroups is by comparing benefits and harms in the same units. In the primary analysis, the number of ILD deaths caused by T-DXd was approximately 13 per 1,000 patients across all subgroups, with 95% UIs spanning approximately 2 to 39. Because this harm does not diminish as recurrence risk decreases, and its uncertainty remains wide, it weighs most heavily in the subgroups where projected benefit is smallest. Meanwhile, the number of breast cancer deaths averted increased roughly sevenfold with increasing residual disease volume, from 16 per 1,000 patients with RCB-I ER-positive disease to 113 per 1,000 patients with RCB-III ER-positive disease. The fixed ILD harm alongside the sharply increasing benefit explains the observed subgroup differences. For patients with very low recurrence risk, the absolute reduction in recurrences with T-DXd may be modest while the risk of treatment-related toxicity remains. Our findings suggest that the T-DXd benefit for ER-positive patients with very low volumes of residual disease is small and sensitive to fatal ILD assumptions. This is without even considering nonfatal toxicity burden, quality of life, and cost. Thus, our data support individualized recurrence-risk assessment rather than routine escalation in patients with low volumes of residual disease.^5^

T-DXd has emerged as an effective escalation option in HER2-positive early breast cancer. In the neoadjuvant setting, DESTINY-Breast11 showed that incorporating sequential T-DXd followed by taxane, trastuzumab, and pertuzumab (THP) in high-risk patients (≥cT3cN0 or cT0– 4cN1–3) improved the pCR rate over dose-dense anthracycline-cyclophosphamide (ddAC) followed by THP (67.3% versus 56.3%).^24^ Because the adjuvant DESTINY-Breast05 trial only enrolled patients who received conventional neoadjuvant chemo-HER2 therapy, how to optimally sequence the adjuvant therapy when neoadjuvant T-DXd is used is unclear. This uncertainty is further increased by the immature follow-up data from DESTINY-Breast11 and the known limitations of pCR as a surrogate for long-term outcomes at the cohort level.^25^ Both studies, as well as the broader HER2 literature, continue to support a risk-adapted approach to treatment, and our study extends this framework by providing modeled estimates to inform clinical decision-making. Incorporation of ctDNA dynamics, functional imaging, and shorter-course regimens may eventually allow escalation to be targeted more precisely than residual disease volume alone permits.^5,7^ Until such data are available, quantifying the expected benefit and harm across the spectrum of residual disease burden can inform treatment decision-making in patients for whom no direct randomized evidence exists.

Our analysis has several limitations. We extrapolated 3-year endpoints from DESTINY-Breast05 to project 10-year clinical outcomes and assumed that T-DXd’s relative benefit extends to lower-volume residual disease, despite limited evidence; however, these assumptions are consistent with the durable and stable treatment effects observed in prior adjuvant trials in HER2-positive breast cancer.^1,26^ We assumed proportional hazards, which may be less appropriate for late events in ER-positive disease.^12^ Our model did not capture nonfatal toxicity burden and quality of life, which could shift the net-benefit threshold. As recurrence risk was only varied across RCB class and ER status, we could not assess variation in benefit across nodal size, tumor stage, or age. Despite these limitations, we thoroughly tested the robustness of our findings. Beyond the trial follow-up, we made assumptions that favored the existing standard of care, T-DM1, allowing T-DXd’s benefit to wane after year 3, modeling more fatal ILD than observed in DESTINY-Breast05, and examining edge cases in which T-DXd benefit was attenuated or any-grade ILD was more common. Despite these assumptions favoring T-DM1, there remained clear 10-year survival gains with T-DXd for RCB-II and RCB-III subgroups. Specifically, these assumptions are conservative for the conclusion that T-DXd benefits patients with RCB-II and RCB-III disease, but not for RCB-I. The small projected gain in RCB-I ER-positive disease depends on fatal ILD assumptions.

In conclusion, our findings support T-DXd for most patients with RCB-II or RCB-III residual disease, while benefits are sensitive to fatal ILD assumptions in RCB-I, particularly in ER-positive tumors. Clinicians may combine these estimates with individualized recurrence-risk assessment and shared decision-making about the trade-off between recurrence reduction and ILD risk when considering T-DXd for residual HER2-positive early breast cancer.

## Data Availability

Sources for model inputs are available in Table 1 and the Supplementary Materials (Table S1). The code used for the analysis can be found on GitHub at the following link: https://github.com/janefan7/Adjuvant-TDXd-Manuscript

## Acknowledgments

None

