## Supplementary Materials for "Adjuvant Trastuzumab Deruxtecan Versus Trastuzumab Emtansine by Residual Cancer Burden in HER2-Positive Early Breast Cancer: A Decision Analysis"

**Supplementary Methods**

Model Calibration

We used logit-normal likelihood functions based on the 3-year overall survival (OS), invasive disease-free survival (iDFS), and distant recurrence-free interval (DRFI) endpoints and approximate proportion of recurrences that were locoregional from the DESTINY-Breast05 trial.^1^ Internal validation of the model fit after calibration was performed using 1,000 parameter sets from the joint posterior distribution obtained from the Incremental Mixture Importance Sampling algorithm.^2^ The posterior draws were used to generate model-predicted 3-year OS, iDFS, and DRFI for both T-DXd and T-DM1 arms, which were subsequently compared against the 3-year trial endpoints (**Figure S1**).

Recurrence-free to Locoregional Recurrence Hazard

To derive the transition hazard from recurrence-free to locoregional recurrence, we used the formula

$\lambda_{RF to LRR} = \frac{prop_{LRR}}{1 - prop_{LRR}} \lambda_{RF to DR}$,

where $prop_{LRR}$ is the calibrated proportion of recurrences that are locoregional and $\lambda_{RF to DR}$is the calibrated transition hazard from recurrence-free to distant recurrence. As such, $1-prop_{LRR}$ is the proportion of recurrences that are distant, and $\frac{prop_{LRR}}{1 - prop_{LRR}}$ represents the odds of locoregional recurrence relative to distant recurrence.

Residual Cancer Burden (RCB)- and Estrogen Receptor (ER)-Specific Recurrence Hazards

To derive RCB- and ER-specific transition hazards for recurrence-free to locoregional recurrence and recurrence-free to distant recurrence, we first transformed the RCB- and ER-specific 5-year event-free survival (EFS) estimates reported in Table S2 ("Received Neoadjuvant HER2-targeted therapy”) of Yau et al^3^ to cumulative hazards of recurrence (locoregional or distant) or death. Since the Yau et al estimates predated the use of T-DM1, we subsequently multiplied by the hazard ratio for invasive disease or death for T-DM1 versus trastuzumab, using the final analysis of iDFS from the KATHERINE trial (median follow-up of 8.4 years).^4^ These quantities were converted to 5-year probabilities to obtain the T-DM1-adjusted 5-year EFS estimates using the formula

$1-exp(-{HR}_{KATHERINE} \times[-log(EFS_{5y})])$, which simplifies to

$1-(EF{S_{5y})}^{{HR}_{KATHERINE}}$,

where ${HR}_{KATHERINE}$ denotes the hazard ratio from KATHERINE

For each RCB/ER subgroup, we then used a numerical root-finding algorithm to estimate a subgroup-specific recurrence multiplier, applied to both the calibration-derived recurrence-free to locoregional recurrence hazard and calibrated T-DM1 recurrence-free to distant recurrence hazard, that most closely reproduced the corresponding T-DM1-adjusted 5-year EFS (**Table S1**). Applying a common multiplier to both locoregional and distant recurrence hazards was done to preserve their calibrated ratio. The calibrated T-DXd hazard ratio for distant recurrence was then applied to the RCB- and ER-specific T-DM1 recurrence-free to distant recurrence hazards to obtain the corresponding hazards under T-DXd.

Locoregional to Distant Recurrence Hazard

As locoregional events were rare in DESTINY-Breast05 and external data on the progression from locoregional to distant recurrence in HER2-positive early breast cancer are limited, we did not estimate a subgroup- or treatment-specific locoregional to distant recurrence hazard. Instead, we calibrated a multiplier with a Uniform(1,2) prior relative to the calibrated T-DM1 recurrence-free to distant recurrence hazard. We then applied this multiplier to the recurrence-free to distant recurrence hazard of the highest-risk subgroup under T-DM1 (i.e., RCB-III ER-positive) and used the resulting absolute hazard as the common locoregional to distant recurrence hazard across both treatment arms and all RCB/ER subgroups. Anchoring on the highest-risk subgroup ensures the locoregional to distant recurrence hazard exceeds the recurrence-free to distant recurrence hazard in every subgroup and treatment arm, preserving a clinically coherent ordering in which locoregional recurrence confers a higher subsequent risk of distant recurrence than does remaining recurrence-free. Because this hazard is shared across arms while the recurrence-free to distant recurrence hazard is lower for T-DXd (given the assumption that T-DXd reduces the recurrence-free to distant recurrence hazard), the specification assumes no residual T-DXd benefit on distant recurrence after a locoregional event and is therefore conservative with respect to T-DXd. The multiplier was weakly identified by the 3-year calibration targets and was retained primarily to propagate uncertainty in post-locoregional recurrence progression.

Treatment Effect Following Interstitial Lung Disease (ILD) Discontinuation

Patients discontinuing T-DXd during the 10-month adjuvant treatment period received an *a priori-*specified fraction of the treatment effect on the log-hazard ratio scale based on the month of discontinuation. The fractions corresponding to months 1 through 10, respectively, were 0.28, 0.49, 0.64, 0.75, 0.83, 0.89, 0.94, 0.97, 0.99, and 1.00. The retained treatment benefit following discontinuation during month *m* was calculated as

$HR_{m}=H{R_{full}}^{b_{m}}$,

where$HR_{full}$denotes the full treatment benefit from T-DXd and $b_{m}$ denotes the fraction of benefit retained for patients who discontinued T-DXd during month *m*. Given that T-DM1 is the current standard of care for HER2-positive early breast cancer and ILD-related discontinuation from T-DM1 was uncommon in DESTINY-Breast05, we assumed that the RCB- and ER-specific T-DM1 recurrence-free to distant recurrence hazards remained unchanged following T-DM1 discontinuation.

ILD Discontinuation and Fatal Hazards

Treatment-specific ILD discontinuation hazards and T-DXd-associated fatal ILD hazards were derived from DESTINY-Breast05, where grades 2-4 ILD indicated discontinuation and grade 5 was fatal. We assumed that all ILD events occurred over the 10-month adjuvant treatment period. Specifically, we first converted the cumulative treatment-specific probability of any-grade ILD over the 10-month period, *P(any ILD)*, to a constant monthly hazard using the formula:

$$\lambda_{any ILD} = \frac{-log[1-P(any ILD)]}{10}$$

As T-DXd-associated fatal ILD risk reported in DESTINY-Breast05 may underestimate real-world ILD toxicity,^5^ we set the probability of fatal ILD among patients assigned to T-DXd to 1.0% in the deterministic threshold analyses, rather than the trial-observed value of 0.25% used during model calibration. We obtained the fatal proportion among any-grade ILD cases with T-DXd using the equation:

$P\left( fatal ILD \right| any ILD) =\frac{P(fatal ILD among patients assigned to T-DXd)}{P(any-grade ILD with T-DXd)}$

We subsequently partitioned the monthly any-grade ILD hazard for T-DXd into fatal and nonfatal ILD hazards:

$$\lambda_{fatal ILD} =\lambda_{any ILD} \times P(fatal ILD | any ILD)$$

$$\lambda_{nonfatal ILD} =\lambda_{any ILD} \times[1 - P(fatal ILD | any ILD)]$$

For T-DM1, the nonfatal ILD hazard was equal to the any-grade ILD hazard since we did not model fatal ILD cases for patients assigned to T-DM1.

Since ILD discontinuation was modeled as a subset of nonfatal ILD, we calculated the treatment-specific monthly ILD discontinuation hazard as follows:

$\lambda_{ILD discontinuation}=\lambda_{nonfatal ILD} \times P(discontinuation | nonfatal ILD)$,

where $P(discontinuation | nonfatal ILD)$ was obtained by dividing the treatment-specific number of ILD discontinuations by the treatment-specific number of nonfatal ILD cases in DESTINY-Breast05.

For the ILD-related probabilities described above, the reference values, uncertainty intervals, distributions for the probabilistic sensitivity analysis, and sources used are presented in **Table S1**. The treatment-specific probability of ILD discontinuation among nonfatal ILD cases was treated as a fixed parameter.

Probabilistic Sensitivity Analysis

For calibrated parameters, we used the posterior mean as the reference value and the 95% credible interval obtained from the posterior distribution as the range. We sampled any-grade ILD probabilities for T-DXd from a logit-normal distribution parameterized using the reference value of 9.6% from DESTINY-Breast05, as well as lower and upper bounds of roughly 5.0% and 21.0% informed by Powell et al^6^ and clinical expertise. This yielded a 95% uncertainty interval of 4.5% to 19.3%. The any-grade ILD probabilities for T-DM1 were sampled from a beta distribution parameterized using event counts from DESTINY-Breast05 and a uniform Beta(1,1) prior. For the primary analysis assuming a more conservative fatal ILD risk, we sampled the probability of fatal ILD among patients assigned to T-DXd from a logit-normal distribution centered at 1.0% and using lower and upper bounds of 0.2% and 3.0% informed by Powell et al,^6^ Park et al,^7^ and clinical expertise. This yielded a 95% uncertainty interval of 0.3% to 3.8%. For the scenario analysis of fatal ILD risk, we sampled the probability of fatal ILD among patients assigned to T-DXd from a beta distribution parameterized using event counts from DESTINY-Breast05 and a uniform Beta(1,1) prior.

**Supplementary Figures and Tables**

**Figure S1. Internal Validation Plot for Model Calibration^a^**

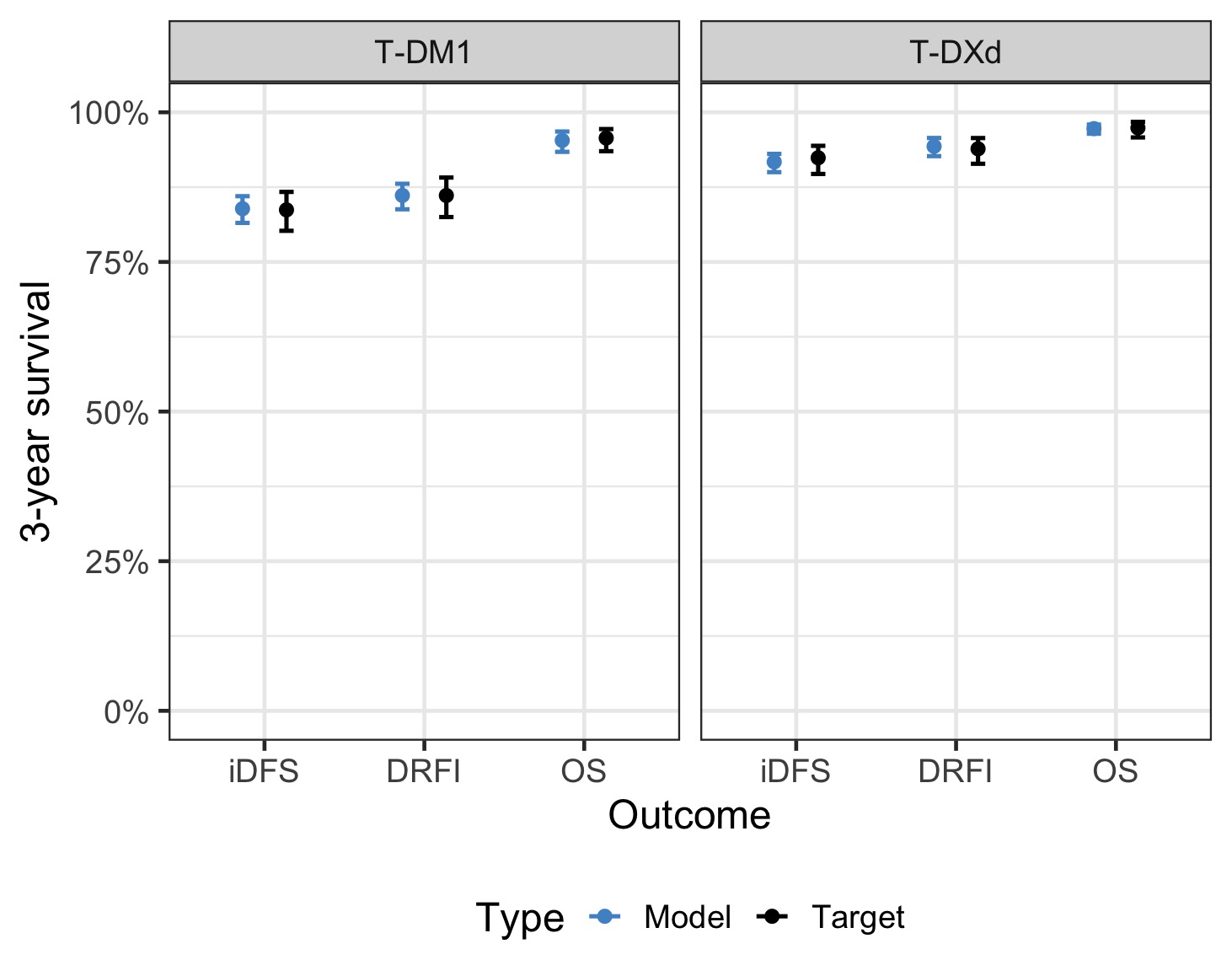

Abbreviations: T-DM1, trastuzumab emtansine; T-DXd, trastuzumab deruxtecan; iDFS, invasive disease-free survival; DRFI, distant recurrence-free interval; OS, overall survival.

^a^ Mean model-predicted outputs are shown as blue circles, along with the corresponding error bars. The 3-year iDFS, DRFI, and OS endpoints from the DESTINY-Breast05 trial are shown as black circles, with the corresponding error bars.

**Figure S2. Prior vs Posterior Distributions of Calibrated Parameters from the IMIS Algorithm**

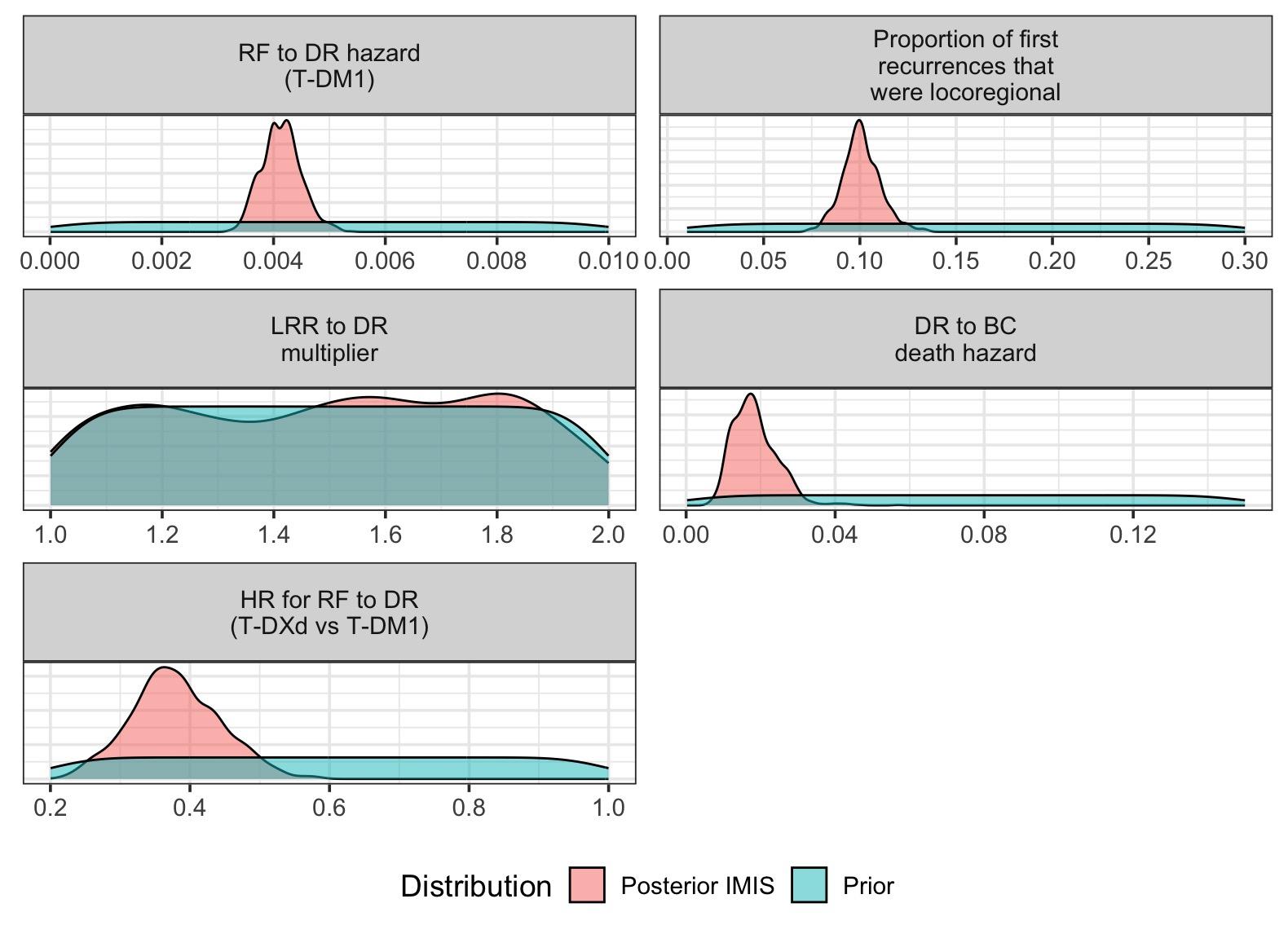

Abbreviations: IMIS, Incremental Mixture Importance Sampling; RF, recurrence-free; DR, distant recurrence; T-DM1, trastuzumab emtansine; LRR, locoregional recurrence; BC, breast cancer; HR, hazard ratio; T-DXd, trastuzumab deruxtecan.

**Figure S3. Marginal Posterior Distributions, Pairwise Scatterplots, and Correlations of Calibrated Parameters from the IMIS Algorithm**

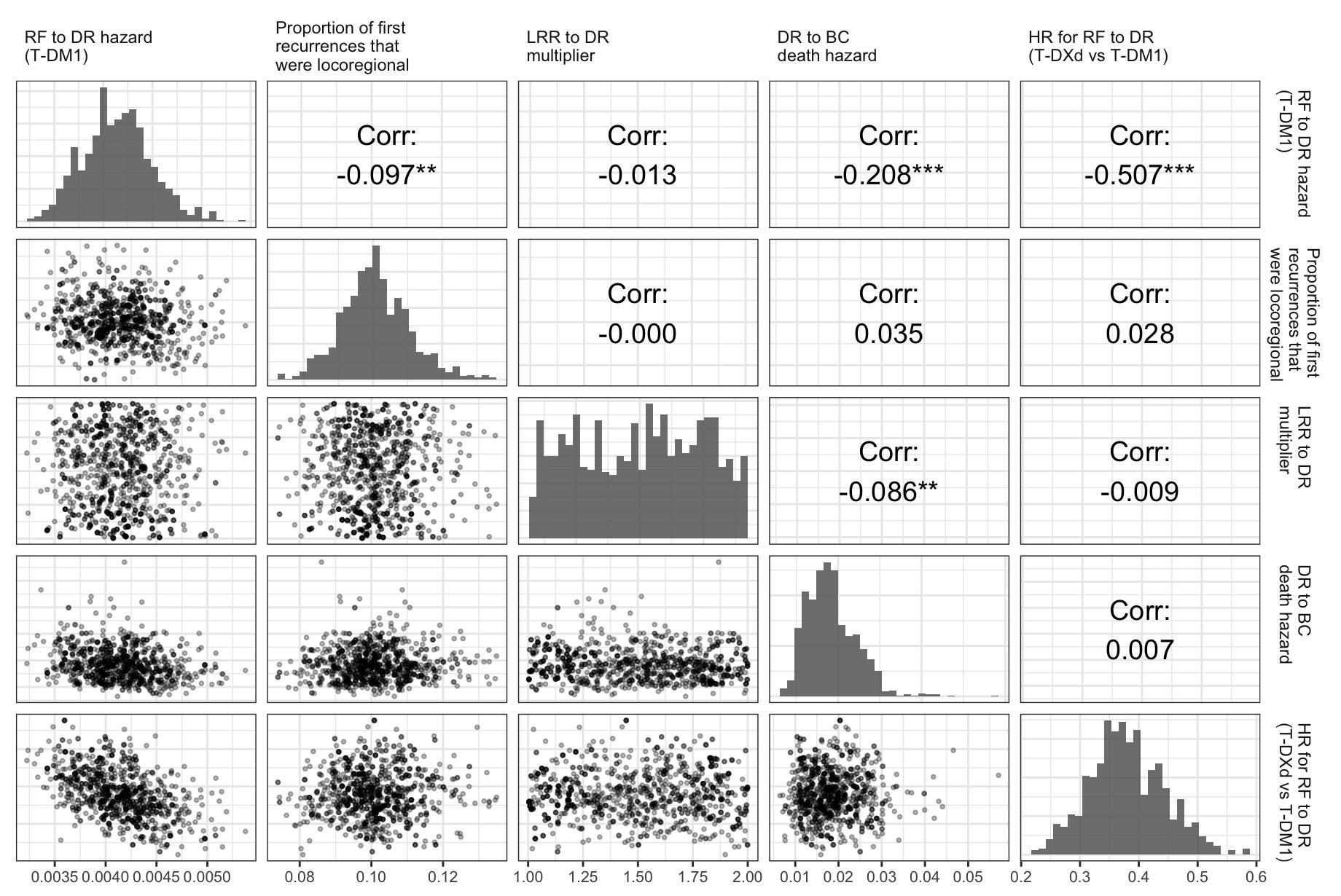

Abbreviations: IMIS, Incremental Mixture Importance Sampling; RF, recurrence-free; DR, distant recurrence; T-DM1, trastuzumab emtansine; LRR, locoregional recurrence; BC, breast cancer; HR, hazard ratio; T-DXd, trastuzumab deruxtecan.

**Table S1. Disease Progression, Mortality, and ILD Parameters for the Clinical Decision-Analytic Model^a^**

| **Model input** | **Reference value^b^** | **Range** | **Distribution for probabilistic sensitivity analysis** | **Source** |
| --- | --- | --- | --- | --- |
| **Disease progression and mortality** | | | | |
| T-DM1 recurrence-free to distant recurrence monthly hazard^c^ | 0.004 | 95% CrI: (0.0035, 0.0049) | Empirically calibrated posterior distribution | Model calibration to the DESTINY-Breast05 trial^1^ |
| Hazard ratio for distant recurrence hazard, T-DXd vs. T-DM1^d^ | 0.380 | 95% CrI: (0.260, 0.509) | Empirically calibrated posterior distribution | Model calibration to the DESTINY-Breast05 trial^1^ |
| Distant recurrence to breast cancer death monthly hazard | 0.019 | 95% CrI: (0.010, 0.030) | Empirically calibrated posterior distribution | Model calibration to the DESTINY-Breast05 trial^1^ |
| Proportion of recurrences that were locoregional | 0.101 | 95% CrI: (0.082, 0.123) | Empirically calibrated posterior distribution | Model calibration to the DESTINY-Breast05 trial^1^ |
| Recurrence-free to locoregional recurrence monthly hazard^c^ | 0.0005 | 95% CrI: (0.0003, 0.0006) | Empirical distribution from joint posterior draws | Derived using calibrated T-DM1 recurrence-free to distant recurrence hazard and calibrated proportion of recurrences that were locoregional |
| Locoregional to distant recurrence multiplier | 1.50 | 95% CrI: (1.02, 1.99) | Empirically calibrated posterior distribution | Model calibration to the DESTINY-Breast05 trial^1^ (weakly identified) |
| Locoregional to distant recurrence monthly hazard | 0.007 | 95% CrI: (0.005, 0.010) | Empirical distribution from joint posterior draws | Derived by applying the calibrated locoregional to distant recurrence multiplier to the recurrence-free to distant recurrence hazard of the highest risk subgroup (RCB-III ER-positive) under T-DM1 |
| Estimated T-DM1-adjusted 5-year EFS by RCB class and ER status^e^ |  | Not varied | – | Derived using Yau et al^3^ and the KATHERINE trial^4^ |
| ER-positive / HER2-positive |  |  |  |  |
| RCB-I | 95.0% |  |  |  |
| RCB-II | 86.2% |  |  |  |
| RCB-III | 71.7% |  |  |  |
| ER-negative / HER2-positive |  |  |  |  |
| RCB-I | 91.6% |  |  |  |
| RCB-II | 77.9% |  |  |  |
| RCB-III | 75.9% |  |  |  |
| RCB- and ER-specific recurrence multiplier |  | Not varied | – | Derived to reproduce the corresponding RCB- and ER-specific T-DM1-adjusted 5-year EFS estimates |
| ER-positive / HER2-positive | | | | |
| RCB-I | 0.120 |  |  |  |
| RCB-II | 0.472 |  |  |  |
| RCB-III | 1.139 |  |  |  |
| ER-negative / HER2-positive |  |  |  |  |
| RCB-I | 0.252 |  |  |  |
| RCB-II | 0.839 |  |  |  |
| RCB-III | 0.933 |  |  |  |
| Background mortality | Age-specific | Not varied | – | 2023 CDC life table for women^8^ |
| **ILD probabilities over 10-month treatment period** | | | | |
| Any-grade ILD, T-DXd | 9.6% | 95% UI: (4.51, 19.27) | Logit-normal(μ =  -2.24, σ = 0.41) | DESTINY-Breast05 trial,^1^ Powell et al,^6^ and clinical expertise |
| Any-grade ILD, T-DM1 | 1.6% | 95% UI: (0.96, 2.76) | Beta(14, 789) | DESTINY-Breast05 trial^1^ |
| ILD discontinuation among nonfatal ILD, T-DXd | 78.7% | Not varied | – | Derived using the number of T-DXd-associated ILD discontinuations and the number of nonfatal ILD cases for T-DXd from the DESTINY-Breast05 trial^1^ |
| ILD discontinuation among nonfatal ILD, T-DM1^f^ | 38.5% | Not varied | – | Derived using the number of T-DM1-associated ILD discontinuations and the number of nonfatal ILD cases for T-DM1 from the DESTINY-Breast05 trial^1^ |
| Fatal ILD among patients assigned to T-DXd^g^ | 1.0% for primary analysis | 95% UI: (0.26, 3.82) for primary analysis; (0.08, 0.89) for scenario analysis | Logit-normal(μ =  -4.60, σ = 0.70) for primary analysis; Beta(3, 805) for scenario analysis | DESTINY-Breast05 trial,^1^ Powell et al,^6^ Park et al,^7^ and clinical expertise |
| Fatal ILD among any-grade ILD, T-DXd | 10.4% for primary analysis | 95% UI: (2.05, 54.66) for primary analysis; (0.66, 13.18) for scenario analysis | Derived from sampled inputs | Derived using the any-grade ILD probability for T-DXd from the DESTINY-Breast05 trial^1^ and fatal ILD probability among patients assigned to T-DXd |

Abbreviations: ILD, interstitial lung disease; CrI, credible interval; RCB, residual cancer burden; ER, estrogen receptor; T-DM1, trastuzumab emtansine; EFS, event-free survival; HER2, human epidermal growth factor receptor 2; T-DXd, trastuzumab deruxtecan; CDC, Centers for Disease Control and Prevention; UI, uncertainty interval.

^a^ Reference values, uncertainty ranges, probability distributions, and data sources used in the model are shown. Credible intervals summarize posterior distributions for calibrated parameters and posterior-derived quantities; uncertainty intervals represent uncertainty in other probabilistic model parameters. See the Supplementary Methods for a detailed description of the parameters. For non-calibrated and non-posterior-derived parameters with specified distributions, uncertainty intervals were obtained using the theoretical 2.5^th^ and 97.5^th^ percentiles of the distributions; for non-calibrated and non-posterior-derived parameters whose distributions were derived using those of other sampled inputs, uncertainty intervals were obtained empirically from probabilistic sensitivity analysis draws.

^b^ For calibrated and posterior-derived quantities, reference values are the posterior means and were used in the deterministic threshold analysis only. Reference values for ILD probabilities were used in the deterministic threshold analysis.

^c^ RCB- and ER-specific hazards for the recurrence-free to locoregional recurrence transition and the T-DM1 recurrence-free to distant recurrence transition were obtained by applying a subgroup-specific recurrence multiplier to both the calibrated recurrence-free to locoregional recurrence hazard and T-DM1 recurrence-free to distant recurrence hazard.

^d^ As T-DXd was assumed to affect the hazard of distant recurrence, the treatment-effect hazard ratio for T-DXd relative to T-DM1 was applied to the RCB- and ER-specific T-DM1 recurrence-free to distant recurrence hazards.

^e^ Used to define recurrence risk by RCB class and ER status. Estimated by using the RCB- and ER-specific 5-year EFS estimates from Table S2 ("Received Neoadjuvant HER2-targeted therapy”) of Yau et al, which predated the use of T-DM1, and adjusting with the iDFS hazard ratio for T-DM1 versus trastuzumab from the final analysis of the KATHERINE trial. Additional derivation details can be found in the Supplementary Methods.

^f^ We did not model fatal ILD for T-DM1, given that no fatal ILD cases were reported for T-DM1 in DESTINY-Breast05. Thus, for T-DM1, the number of nonfatal ILD cases was equal to the number of any-grade ILD cases.

^g^ To account for the DESTINY-Breast05 trial’s potential underestimation of real-world ILD toxicity, we used a conservative value of 1.0% in the deterministic threshold analysis and in the parameterization of the distribution used in the primary probabilistic sensitivity analysis, informed by pooled T-DXd studies and clinical expertise. The trial-observed value of 0.25% was used in calibrating the model to the 3-year trial endpoints and informed the parameterization of the probabilistic distribution used in the scenario analysis of fatal ILD risk; a deterministic threshold analysis was not conducted as part of the scenario analysis.

**Table S2. Mean Differences in 10-Year Overall Survival with T-DXd vs T-DM1 Under Alternative Treatment Effect Scenarios^a^**

| **Treatment effect scenario** | **RCB-I** | **RCB-II** | **RCB-III** |
| --- | --- | --- | --- |
| **ER-positive / HER2-positive** | | | |
| Effect ends after year 3 | -0.4  (-2.7 to 0.7) | 1.8  (-0.5 to 3.3) | 4.7  (2.3 to 6.9) |
| Persistent effect through year 10 | 0.7  (-1.6 to 2.2) | 5.8  (2.6 to 9.2) | 12.9  (7.5 to 19.1) |
| **ER-negative / HER2-positive** | | | |
| Effect ends after year 3 | 0.5  (-1.8 to 1.7) | 3.6  (1.1 to 5.5) | 4.0  (1.4 to 6.0) |
| Persistent effect through year 10 | 2.7  (0.1 to 4.9) | 10.0  (5.6 to 15.1) | 11.0  (6.2 to 16.4) |

Abbreviations: T-DXd, trastuzumab deruxtecan; T-DM1, trastuzumab emtansine; RCB, residual cancer burden; ER, estrogen receptor; HER2, human epidermal growth factor receptor 2.

^a^ Presented values are the means and corresponding 95% uncertainty intervals. Positive values favor T-DXd while negative values favor T-DM1 for the estimated improvement in 10-year OS, under a conservative fatal ILD risk and two alternative treatment effect scenarios: (i) immediate cessation of the treatment effect after year 3 (hazard ratio = 1); and (ii) persistence of the treatment effect through year 10. Patients who completed T-DXd treatment received the full hazard ratio through year 3. Those who discontinued T-DXd due to ILD during the 10-month treatment period retained partial benefit based on the month of discontinuation through year 3.

**Figure S4. Probabilistic Distribution of the 10-Year Overall Survival Difference with T-DXd vs T-DM1 Under the 3-Year Treatment Effect Scenario^a^**

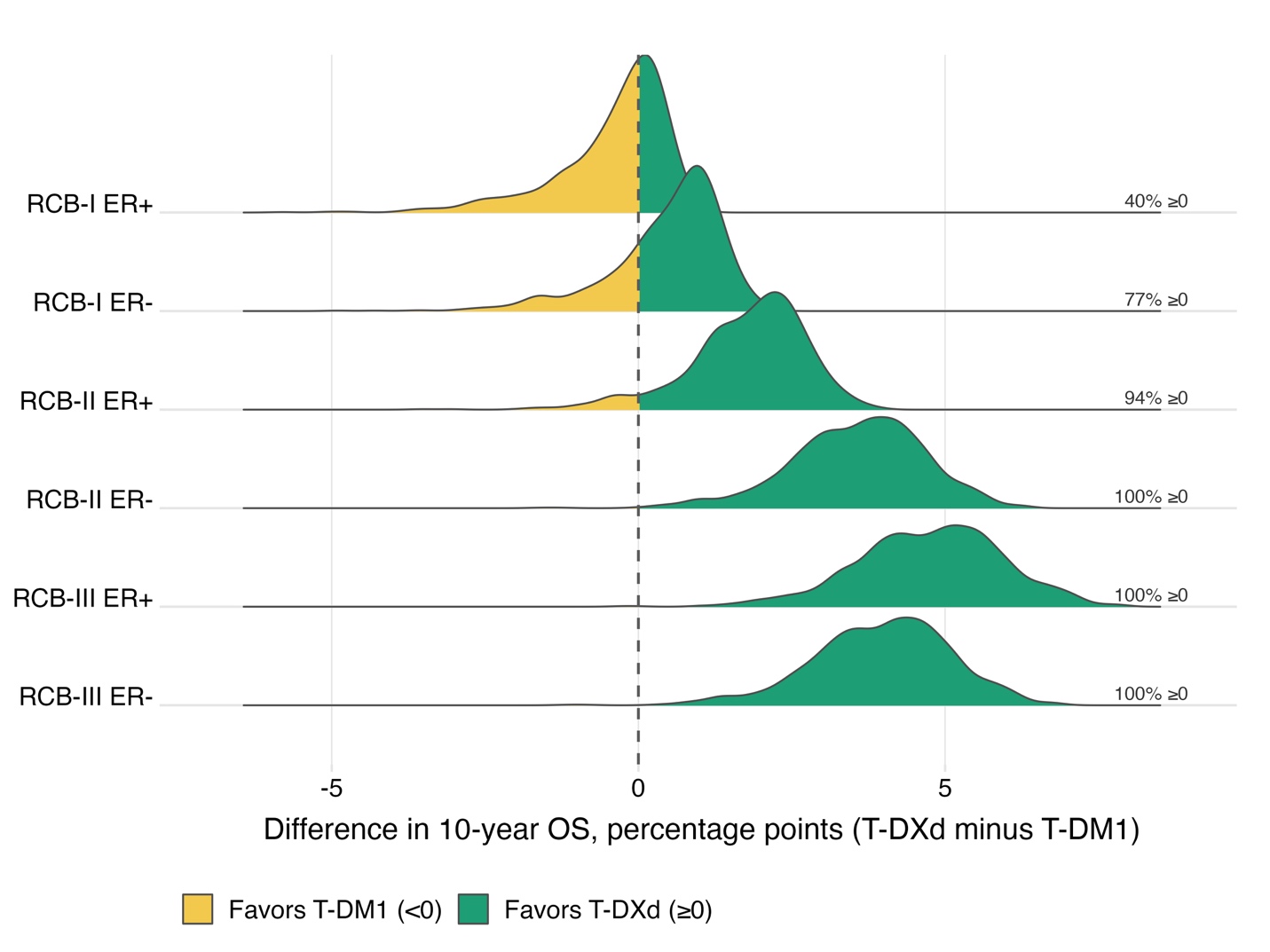

Abbreviations: T-DXd, trastuzumab deruxtecan; T-DM1, trastuzumab emtansine; RCB, residual cancer burden; ER, estrogen receptor; OS, overall survival.

^a^ Shows the distribution of the absolute differences in 10-year OS with T-DXd vs T-DM1 across 1,000 simulations, reported in percentage points, under a conservative fatal ILD risk and an alternative treatment-effect assumption in which the T-DXd treatment effect was assumed to cease immediately after year 3. Positive values favor T-DXd while negative values favor T-DM1. The percentage of 1,000 simulations favoring T-DXd is shown to the right. Patients who completed T-DXd treatment received the full hazard ratio through year 3. Those who discontinued T-DXd due to ILD during the 10-month treatment period retained partial benefit based on the month of discontinuation through year 3.

**Figure S5. Probabilistic Distribution of the 10-Year Overall Survival Difference with T-DXd vs T-DM1 Under the Persistent Treatment Effect Scenario^a^**

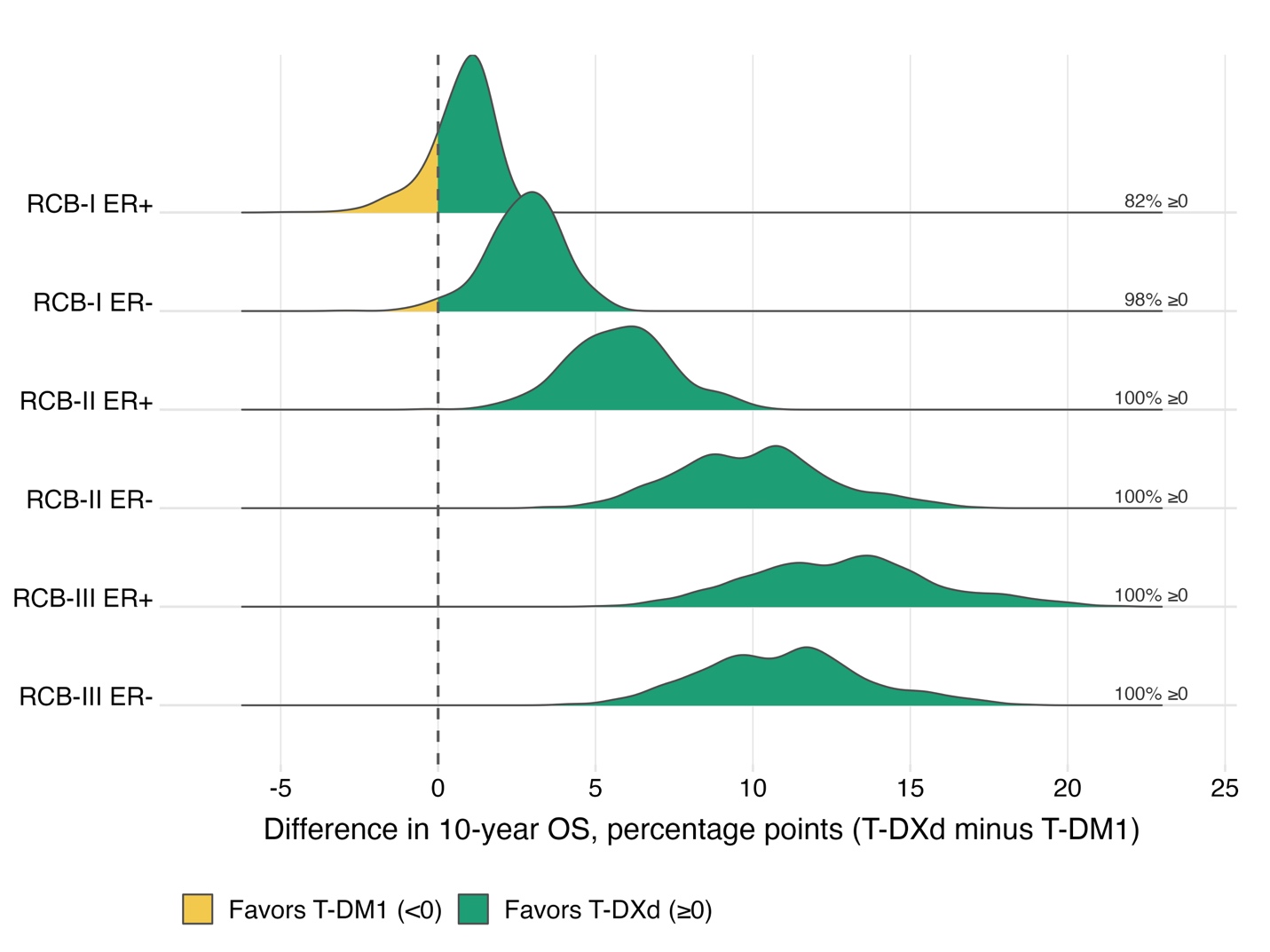

Abbreviations: T-DXd, trastuzumab deruxtecan; T-DM1, trastuzumab emtansine; RCB, residual cancer burden; ER, estrogen receptor; OS, overall survival.

^a^ Shows the distribution of the absolute differences in 10-year OS with T-DXd vs T-DM1 across 1,000 simulations, reported in percentage points, under a conservative fatal ILD risk and an alternative treatment-effect assumption in which the T-DXd treatment effect was assumed to persist through year 10. Positive values favor T-DXd while negative values favor T-DM1. The percentage of 1,000 simulations favoring T-DXd is shown to the right. Patients who completed T-DXd treatment received the full hazard ratio through year 3. Those who discontinued T-DXd due to ILD during the 10-month treatment period retained partial benefit based on the month of discontinuation through year 3.

**Table S3. Mean Differences in 10-Year Overall Survival with T-DXd vs T-DM1 Under DESTINY-Breast05-Informed Fatal ILD Risk and All Treatment Effect Scenarios^a^**

| **Treatment effect scenario** | **RCB-I** | **RCB-II** | **RCB-III** |
| --- | --- | --- | --- |
| **ER-positive / HER2-positive** | | | |
| Linear waning after year 3 until year 10^b^ | 1.2  (0.4 to 1.9) | 5.1  (3.2 to 7.4) | 10.5  (6.8 to 14.7) |
| Effect ends after year 3^c^ | 0.5  (-0.1 to 0.9) | 2.6  (1.6 to 3.7) | 5.5  (3.7 to 7.4) |
| Persistent effect through year 10^c^ | 1.6  (0.8 to 2.6) | 6.6  (4.1 to 9.7) | 13.6  (8.7 to 19.6) |
| **ER-negative / HER2-positive** | | | |
| Linear waning after year 3 until year 10^b^ | 2.7  (1.6 to 4.1) | 8.3  (5.4 to 11.8) | 9.1  (5.8 to 12.8) |
| Effect ends after year 3^c^ | 1.3  (0.6 to 2.1) | 4.3  (2.8 to 6.0) | 4.7  (3.1 to 6.5) |
| Persistent effect through year 10^c^ | 3.6  (2.1 to 5.4) | 10.8  (6.8 to 15.7) | 11.8  (7.5 to 17.0) |

Abbreviations: T-DXd, trastuzumab deruxtecan; T-DM1, trastuzumab emtansine; RCB, residual cancer burden; ER, estrogen receptor; HER2, human epidermal growth factor receptor 2.

^a^ Presented values are the means and corresponding 95% uncertainty intervals, under the scenario analysis informed by the DESTINY-Breast05-observed risk of fatal ILD among patients assigned to T-DXd. Positive values favor T-DXd while negative values favor T-DM1 for the estimated improvement in 10-year OS. Patients who completed T-DXd treatment received the full hazard ratio through year 3. Those who discontinued T-DXd due to ILD during the 10-month treatment period retained partial benefit based on the month of discontinuation through year 3.

^b^ Represents the primary treatment-effect persistence assumption, with the T-DXd treatment effect waning linearly after year 3 until year 10.

^c^ Represents the two alternative treatment effect scenarios: (i) immediate cessation of the treatment effect after year 3 (hazard ratio = 1); and (ii) persistence of the treatment effect through year 10.
